# Implementing ESAMyN: Ecuador’s National Standard for Mother- and Baby-Friendly Health Facilities

**DOI:** 10.64898/2026.07.28.26359056

**Authors:** Betzabé Tello, Marysol Ruilova-Maldonado, Alfredo José Olmedo-Valarezo

## Abstract

**Background:** Ecuador’s Mother- and Baby-Friendly Health Facilities Standard (ESAMyN, *Establecimientos de Salud Amigos de la Madre y el Niño*) is an integrated national quality improvement strategy that combines evidence-based recommendations for antenatal, childbirth, postpartum, newborn and breastfeeding care within a single implementation and certification framework. However, little is known about the factors influencing its implementation in routine health services. This study explored the organizational, contextual and behavioural factors that facilitated or hindered ESAMyN implementation across public health facilities.

**Methods:** We conducted a secondary qualitative analysis of 36 semi-structured interviews originally collected during a programme systematization in 11 public health facilities across five Ecuadorian provinces. Participants included health authorities, managers, ESAMyN focal points, quality coordinators, healthcare professionals and women receiving maternity services. Interview transcripts were analysed inductively using thematic analysis.

**Results:** Implementation was perceived as an organizational transformation rather than the adoption of a new clinical guideline. Strong leadership, multidisciplinary teamwork, continuous training and institutional commitment facilitated implementation and promoted the adoption of respectful, family-centred and evidence-based maternity care practices. However, staff turnover, workload, infrastructure limitations, shortages of equipment and funding, and reliance on local adaptation challenged implementation and contributed to variability across facilities. Participants emphasized that sustaining ESAMyN required institutionalization beyond certification through continuous monitoring, refresher training, supportive supervision and stable organizational support.

**Conclusions:** Integrated national quality standards can strengthen maternal and newborn care through sustained leadership, adequate resources and continuous quality improvement, providing practical lessons for implementing WHO recommendations in routine health services.

## Introduction

High-quality maternal and newborn care requires coordinated, evidence-based interventions throughout pregnancy, childbirth, the immediate postnatal period and early infant feeding (World Health Organization, 2025b; World Health Organization & United Nations Children’s Fund (UNICEF), 2025). Over the past decade, the World Health Organization (WHO) has developed comprehensive recommendations to improve care across this continuum, including guidance on antenatal care for a positive pregnancy experience, intrapartum care for a positive childbirth experience, postnatal care for mothers and newborns, and breastfeeding protection, promotion and support through the Baby-Friendly Hospital Initiative (BFHI) (World Health Organization & United Nations Children’s Fund (UNICEF), 2025). Together, these recommendations promote not only the delivery of effective clinical interventions but also respectful, person-centred care that improves women’s and newborns’ health outcomes and care experiences (World Health Organization & United Nations Children’s Fund (UNICEF), 2025).

Translating these recommendations into routine health services requires more than the adoption of individual clinical guidelines (Wojcieszek et al., 2023; World Health Organization & United Nations Children’s Fund (UNICEF), 2018). It requires health systems to implement integrated models of care that ensure continuity across maternal and newborn services while promoting consistent quality improvement, multidisciplinary collaboration and accountability (World Health Organization, 2025c). However, evidence describing how countries operationalize multiple evidence-based recommendations within a single national quality standard remains limited, particularly regarding the implementation processes that facilitate or hinder their adoption in routine practice (Peven et al., 2021; Vallely et al., 2023).

In Ecuador, the Ministry of Public Health developed the Mother- and Baby-Friendly Health Facilities Standard (Establecimientos de Salud Amigos de la Madre y el Niño, ESAMyN) as part of the national strategy to improve the quality of maternal and newborn care (Ministerio de Salud Pública del Ecuador, 2021). Introduced in 2016, ESAMyN was initially inspired by the principles of the Baby-Friendly Hospital Initiative but progressively evolved into a comprehensive national quality standard for the continuum of maternal and newborn care (Ministerio de Salud Pública del Ecuador, 2016). Guided by Ecuador’s Model of Comprehensive Health Care (Modelo de Atención Integral de Salud, MAIS) and national maternal and newborn health priorities, the updated 2021 standard integrates WHO recommendations together with national clinical guidelines into a single implementation and certification framework for health facilities (Ministerio de Salud Pública del Ecuador, 2021).

The current ESAMyN standard is structured around four interrelated components: a general component, a prenatal care component, a childbirth and postpartum care component, and a breastfeeding component. By integrating these components within a single mandatory national standard, ESAMyN seeks to strengthen continuity of care, improve women’s and newborns’ experiences throughout the maternal care pathway, and promote consistent implementation of evidence-based practices across health facilities (Figure 1)(Ministerio de Salud Pública del Ecuador, 2021).

**Figure 1.**
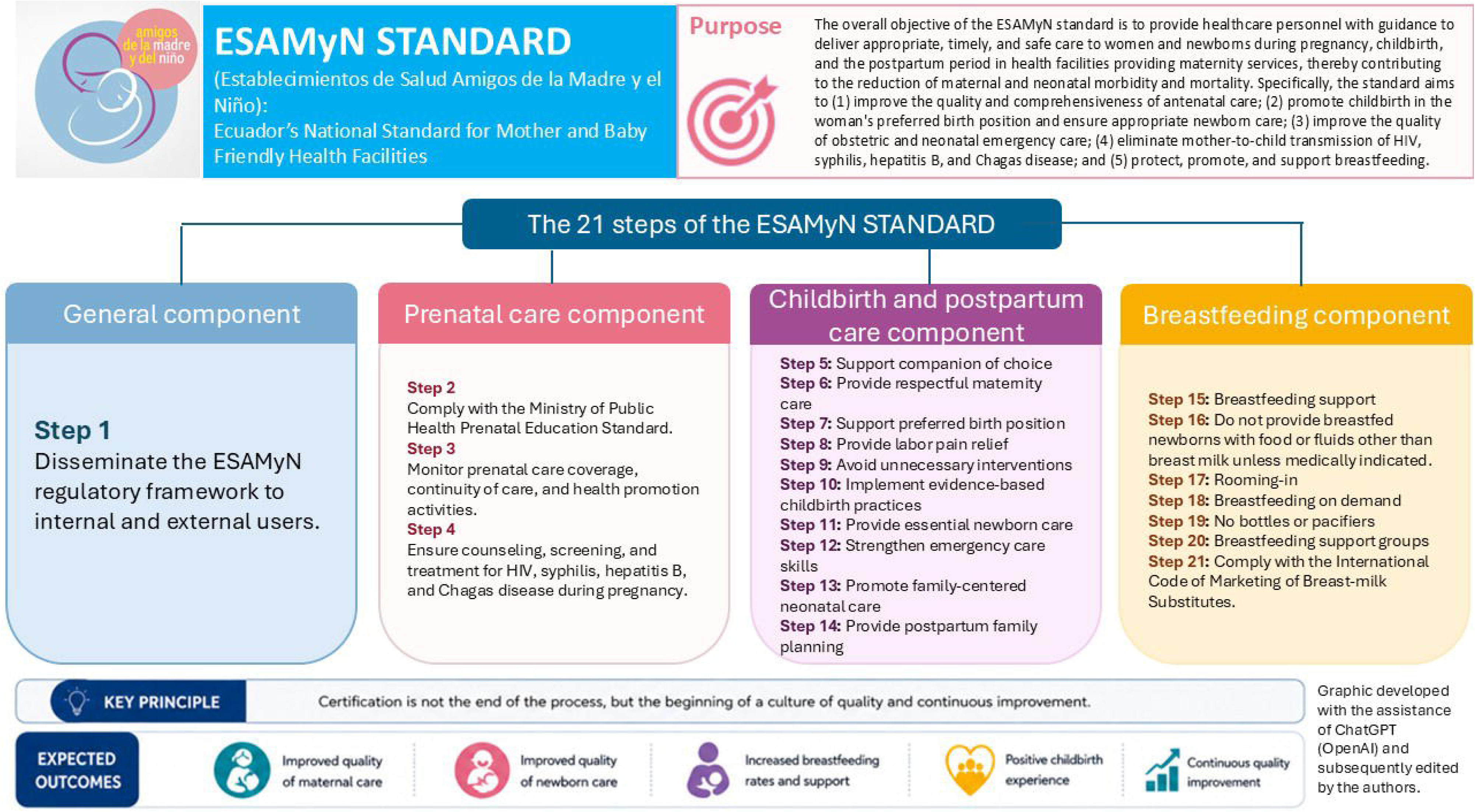
Overview of the ESAMyN Standard for Certification of Mother- and Baby-Friendly Health Facilities in Ecuador.

Although the effectiveness of many individual maternal and newborn interventions is well established, less is known about how integrated national quality standards for the continuum of maternal and newborn care are implemented in routine health services. Understanding the organizational, contextual and behavioural factors that influence implementation is essential for strengthening national quality improvement strategies and may provide lessons for other countries seeking to operationalize WHO recommendations through integrated models of maternal and newborn care.

This study aimed to explore the implementation of Ecuador’s National Standard for Mother- and Baby-Friendly Health Facilities (ESAMyN) and identify the organizational, contextual and behavioural factors that facilitated or hindered its implementation across public health facilities.

## Key messages

Translating WHO recommendations into routine maternal and newborn care requires integrated implementation strategies, yet evidence on how national quality standards are implemented remains scarce.

Ecuador’s ESAMyN standard evolved from the Baby-Friendly Hospital Initiative into an integrated national quality standard for the continuum of maternal and newborn care.

Implementation was enabled by strong leadership, multidisciplinary teamwork and continuous quality improvement, but challenged by staff turnover, resource limitations and infrastructure constraints.

Countries seeking to operationalize WHO maternal and newborn recommendations may benefit from integrated implementation and certification frameworks that strengthen continuity of care across pregnancy, childbirth, postpartum and breastfeeding.

## Methods

### Study design

This study was a secondary qualitative analysis of semi-structured interviews originally conducted as part of a programme systematization commissioned by UNICEF and the Ecuadorian Ministry of Public Health to document the implementation of the National Mother-and-Baby-Friendly Health Facilities Standard (ESAMyN).

### Study setting

The study was conducted in 11 public health facilities implementing ESAMyN across five provinces of Ecuador (Esmeraldas, Manabí, Pichincha, Guayas, Chimborazo). ESAMyN is a national quality improvement standard that integrates evidence-based recommendations for antenatal, childbirth, postpartum and breastfeeding care into a single implementation and certification framework.

### Participants and data collection

Thirty-six semi-structured interviews were conducted between August and September 2025 using purposive sampling. Participants included health authorities, hospital managers, ESAMyN focal points, quality coordinators, physicians, nurses, and women receiving maternity services. Interviews lasted approximately 60 minutes and were conducted by trained researchers using interview guides tailored to each participant group. The interviews explored experiences with ESAMyN implementation, perceived facilitators and barriers, changes in clinical practice, and opportunities for strengthening implementation.

### Data analysis

Interviews were audio-recorded, transcribed verbatim, and anonymized before analysis. Two researchers independently coded the transcripts using an inductive approach. Codes with similar meanings were grouped into broader categories and themes through an iterative process. Differences in coding were resolved through discussion until consensus was reached. The analysis explored facilitators, barriers, implementation strategies, and perceived changes associated with ESAMyN implementation.

### Ethical considerations

The interviews were originally conducted for programme systematization rather than research purposes. This study involved the secondary analysis of anonymized interview transcripts. According to Ecuadorian Ministerial Agreement 00005-2022 (Article 43), secondary analysis of anonymized data is classified as research without risk and does not require review by a Research Ethics Committee.

## Results

### Participant characteristics

Primero incluiría un párrafo corto y una tabla.

Thirty-six participants from 11 public health facilities across five Ecuadorian provinces were interviewed. Participants represented multiple levels of the health system, including national and district health authorities, hospital managers, ESAMyN focal points, quality coordinators, physicians, nurses, and women receiving maternity services. This diversity of perspectives allowed triangulation of implementation experiences across managerial, clinical, and user levels (Table 1).

**Table 1.**
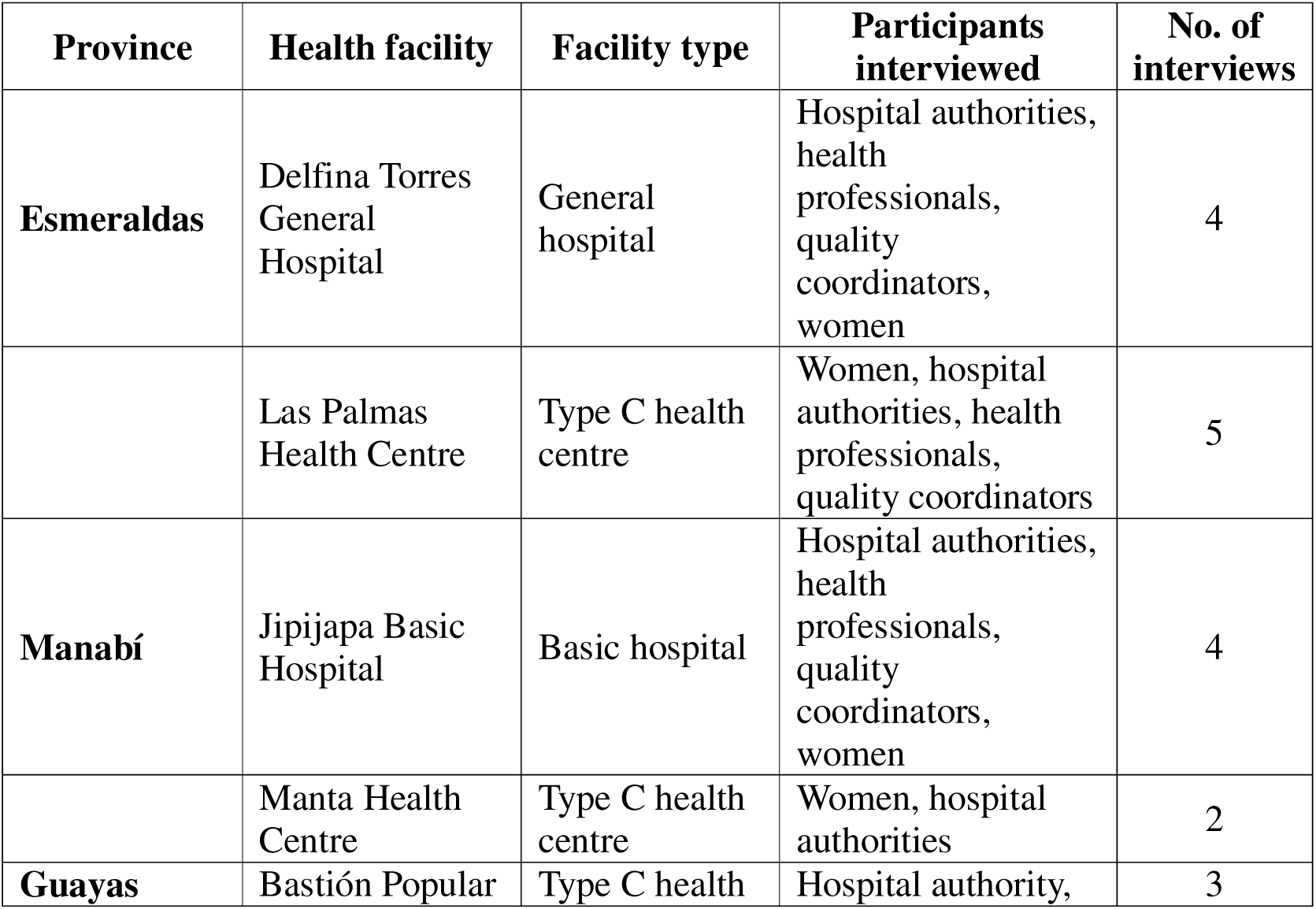
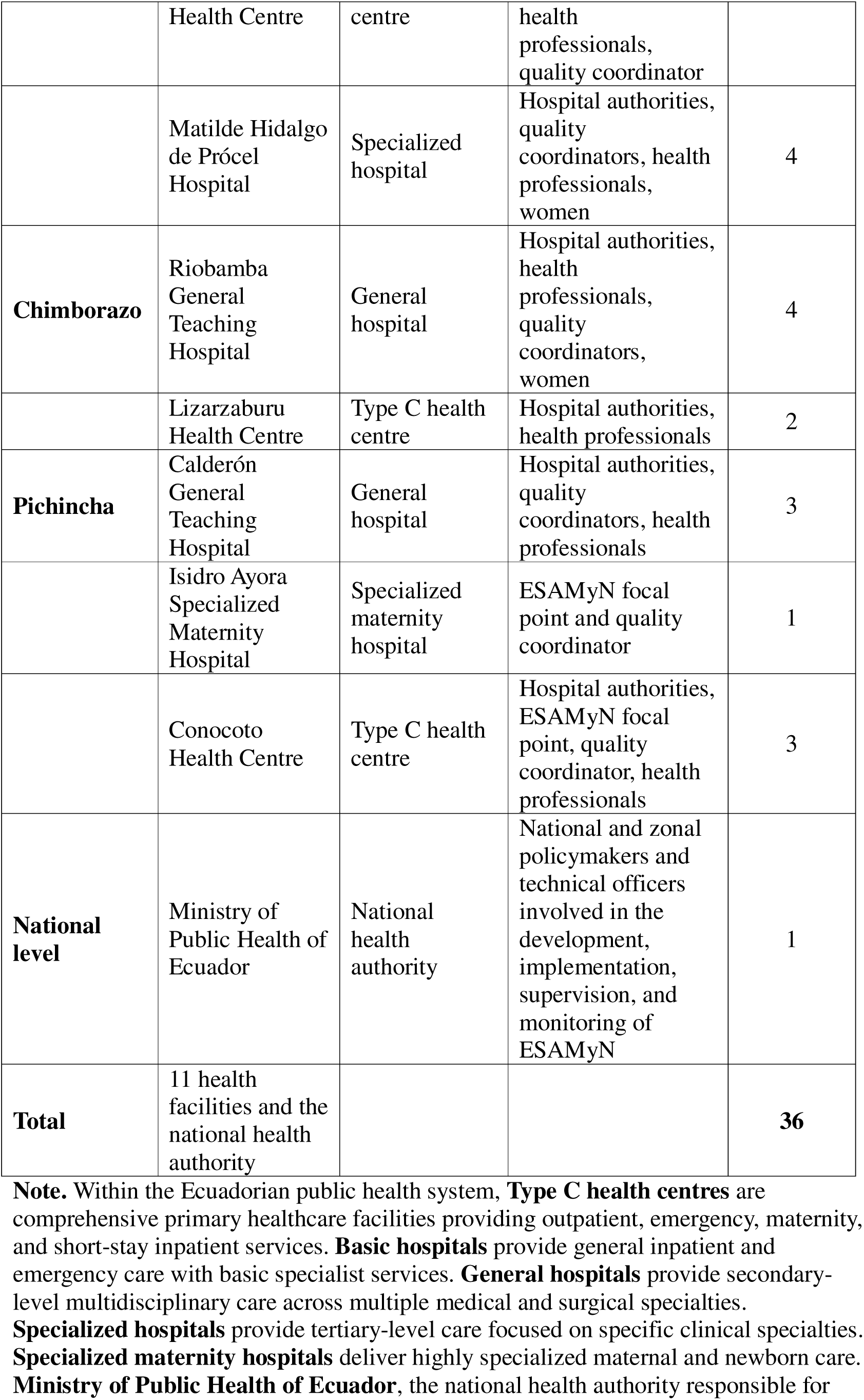
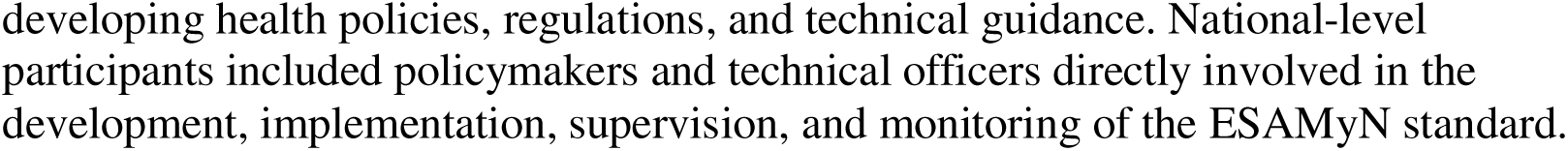
Characteristics of health facilities and participants included in the qualitative study (n = 36 interviews)

### Theme 1. Overcoming resistance to change required strong leadership and institutional commitment

Participants consistently described ESAMyN implementation as a complex organizational change process that initially encountered considerable resistance. Across facilities, changing long-established maternity care practices challenged existing professional routines, particularly those related to birth companionship, women’s participation during childbirth, and the adoption of respectful maternity care practices. Several participants acknowledged that some health professionals questioned the need to modify established ways of working or were reluctant to adopt new approaches.

> *“It was very difficult because we had to change the whole way we attended births, including allowing family members to be present.”* (Quality coordinator)

Resistance was particularly evident during the early stages of implementation and was more frequently attributed to professionals who had worked for many years under traditional models of care. Participants explained that introducing ESAMyN required changing not only clinical practices but also professional attitudes and organizational culture.

To address these challenges, facilities relied on strong institutional leadership. Hospital managers, quality improvement units, ESAMyN focal points, and multidisciplinary committees coordinated implementation, promoted communication across departments, and maintained institutional commitment throughout the certification process. Leadership extended beyond administrative oversight and involved motivating staff, addressing concerns, resolving operational barriers, and reinforcing the importance of the new model of care.

> *“They were the leaders from each service, including medical coordinators and nursing staff.”* (Hospital authority)

In some facilities, participants described how institutional leaders adopted firm positions to ensure compliance with the standard when resistance persisted.

> *“Some physicians said, ‘I don’t want to learn that.’ The manager told them that if they did not agree with the standard, they might need to work somewhere else.”* (Quality coordinator)

Over time, participants reported that resistance gradually diminished as staff became more familiar with the objectives of ESAMyN and observed its benefits in daily practice. Multidisciplinary committees and continuous engagement fostered a sense of shared responsibility, transforming implementation from an externally driven certification process into a collective institutional commitment to improving maternal and newborn care.

### Theme 2. Continuous capacity building strengthened implementation, but sustaining competencies remained challenging

Participants consistently described continuous training as one of the main facilitators of ESAMyN implementation. Beyond the initial certification process, facilities developed ongoing educational activities, including multidisciplinary workshops, clinical simulations, bedside mentoring, and periodic refreshers. These strategies helped standardize clinical practices, reinforce respectful maternity care, and promote shared responsibility across professional groups.

One quality coordinator described how training was deliberately extended beyond maternity staff to strengthen a common understanding of the standard throughout the institution:

> *“Even during training sessions, we included all personnel… obstetric emergencies, neonatal resuscitation…”* (Quality coordinator, Manabí)

Participants emphasized that training promoted organizational learning rather than simply fulfilling certification requirements. Assigning topic leaders, integrating education into routine quality improvement activities, and providing continuous supervision facilitated the incorporation of ESAMyN into everyday clinical practice.

Despite these efforts, participants consistently identified maintaining staff competencies as an ongoing challenge. High staff turnover, rotating residents, competing clinical demands, and limited opportunities for protected training time required facilities to repeat educational activities continuously. Several participants perceived that initial training alone was insufficient to sustain implementation over time.

One participant explained:

> *“I don’t think the training has been sufficient… we need clinical simulation and a sustainability approach.”* (Health professional, Pichincha)

In some facilities, the lack of formal training on newly introduced tools forced professionals to rely on self-directed learning, raising concerns about inconsistent implementation.

> *“They didn’t train us; we trained ourselves.”* (Quality coordinator, Manabí)

Participants therefore viewed capacity building as a continuous organizational process rather than a one-time intervention. However, sustaining staff competencies required ongoing institutional investment to address workforce turnover, workload pressures, and evolving training needs, all of which threatened the long-term consistency of ESAMyN implementation.

### Theme 3. Resource constraints prompted local adaptation and collective action, but limited consistent implementation

Participants across facilities described insufficient funding, inadequate infrastructure, shortages of equipment and supplies, and logistical constraints as persistent barriers to implementing ESAMyN. These limitations affected both hospital-level changes and the capacity of district and zonal teams to provide supervision and technical support. Participants stressed that facilities were expected to comply with the standard even when the necessary physical conditions or resources were unavailable. As one hospital authority explained:

“Sometimes we demand a great deal, but [staff] do not have the tools they need.”

*(Hospital authority, Manabí)*

Infrastructure constraints were especially evident in older facilities and hospitals with limited physical space. Participants described maternity areas that could not easily be modified, insufficient space for prenatal education and breastfeeding support, and environmental conditions that interfered with recommended practices. These limitations sometimes delayed implementation or prevented facilities from meeting all certification requirements. A hospital authority noted:

> “The hospital is 40 years old… it is difficult to have state-of-the-art infrastructure.”
>
> *(Hospital authority, Chimborazo)*

Rather than abandoning implementation, facilities developed locally adapted responses. Participants described reorganizing existing areas, producing their own protocols and monitoring tools, mobilizing community partnerships, and obtaining materials through fundraising activities. In some hospitals, the changes required for ESAMyN were financed almost entirely through collective initiatives led by staff:

> “We organized bingo games and raffles, and sold *tonga* to set up our birthing room.”
>
> *(Quality coordinator, Manabí)*

These forms of self-management extended to basic operating materials. Participants reported conducting community workdays, printing forms at home, and personally paying for educational and communication resources. One focal point stated:

“The humidifiers, roll-up banners, and signage were paid for out of the staff’s own pockets.”

*(ESAMyN focal point and quality coordinator, Pichincha)*

External and intersectoral partnerships partially helped facilities address these gaps. Participants identified support from UNICEF, district and zonal authorities, local governments, and other organizations as an important source of equipment, supplies, training, and infrastructure improvements. In Esmeraldas, a district participant recalled:

“UNICEF supported us… with some supplies and equipment that we did not have.”

*(District participant, Esmeraldas)*

However, access to this support was not uniform across facilities or over time. Some establishments received external resources during particular stages, while others continued to depend largely on local initiative. Logistical limitations were also reported beyond the hospitals themselves. Zonal and district teams sometimes lacked institutional transportation for monitoring visits and training activities:

“We have not had access to transportation… we have had to travel by taxi.”

*(Zonal/district representative, Guayaquil)*

Overall, local creativity, staff commitment, and collaboration with external actors enabled ESAMyN implementation to progress despite substantial resource constraints. Nevertheless, participants’ accounts showed that progress was uneven and frequently depended on extraordinary efforts, including personal financial contributions, that were outside formal institutional financing mechanisms.

### Theme 4. ESAMyN promoted more respectful and family-centred maternity care, although implementation remained inconsistent

Participants consistently perceived that ESAMyN had transformed the organization and delivery of maternal and newborn care by promoting respectful, family-centred, and evidence-based practices. Across facilities, health professionals described a gradual shift away from provider-centred models toward care that recognized women’s rights, encouraged informed decision-making, and supported greater family involvement throughout pregnancy, childbirth, and the postpartum period.

Participants identified several practices that became more common following implementation, including the presence of a birth companion, immediate skin-to-skin contact, delayed umbilical cord clamping, early initiation of breastfeeding, and the use of non-pharmacological methods for pain relief. These changes were perceived as improving both the quality of care and women’s childbirth experiences.

One health professional reflected on this transformation:

> *“Before, everything depended on the professional. Now we understand that the woman and her baby are at the centre of care.” (Health professional)*

Women also described feeling more informed, respected, and involved in decisions regarding their care. Several participants valued receiving explanations during labour, having the opportunity to express their preferences, and being accompanied by a family member throughout the birth process.

> *“They explained everything they were going to do, asked for my consent, and allowed my husband to stay with me. That made me feel calm and safe.” (Woman participant)*

Despite these perceived improvements, participants acknowledged that implementation remained uneven across facilities and among health professionals. Long-standing clinical practices were not always replaced, and adherence to ESAMyN recommendations varied according to staffing, workload, infrastructure, and individual professional commitment. Participants explained that while many professionals had fully adopted the new model of care, others continued to rely on previous routines, resulting in inconsistent experiences for women.

One participant noted:

> *“Some colleagues have embraced the change completely, while others still provide care the way they always have.” (Health professional)*

Women similarly reported variability in their experiences. Although many described respectful care and effective communication, others indicated that the quality of interpersonal care depended on the professional attending them, suggesting that respectful maternity care had not yet been fully institutionalized.

Overall, participants viewed ESAMyN as an important catalyst for improving maternal and newborn care. However, they emphasized that sustaining respectful, evidence-based practices required continuous reinforcement to ensure that improvements were consistently experienced by all women, regardless of facility or provider.

### Theme 5. Sustaining ESAMyN requires institutionalization beyond certification and individual commitment

Participants consistently emphasized that achieving certification represented an important milestone but not the endpoint of implementation. Maintaining ESAMyN standards required continuous leadership, regular monitoring, refresher training, and institutional mechanisms capable of preserving changes despite staff turnover and competing organizational priorities. Several participants expressed concern that, without ongoing reinforcement, previously established practices could gradually decline.

Participants highlighted that sustainability depended on embedding ESAMyN within routine management processes rather than treating it as a time-limited project. Integrating the standard into quality improvement systems, supervisory activities, staff induction, and institutional planning was considered essential for maintaining progress after certification.

> *“Certification is not the end; it is the beginning of maintaining the standards every day.” (Hospital authority)*

However, participants also recognized that sustaining implementation remained challenging because many activities continued to rely on the commitment of a small number of highly motivated individuals. ESAMyN focal points, quality teams, and clinical leaders frequently assumed additional responsibilities to organize training, monitor compliance, and coordinate improvement activities beyond their routine duties.

> *“If the focal point leaves, everything slows down until someone else takes over.” (ESAMyN focal point)*

Participants further explained that long-term sustainability required continued institutional investment in infrastructure, equipment, human resources, and supportive supervision. While external technical assistance had facilitated implementation in many facilities, participants perceived that maintaining the standard increasingly depended on local institutional capacity. They stressed that sustained political and managerial support at national, zonal, district, and facility levels was necessary to avoid losing the progress already achieved.

> *“The commitment has to come from the institution, not only from the people who are enthusiastic about ESAMyN.” (Quality coordinator)*

Despite these challenges, participants remained optimistic about the future of ESAMyN. They perceived that the implementation process had generated lasting organizational changes, strengthened multidisciplinary collaboration, and increased awareness of respectful maternal and newborn care. Nevertheless, they agreed that consolidating these achievements would require moving from implementation driven by individual commitment toward implementation supported by stable institutional systems.

## Discussion

This study provides one of the first qualitative analyses of the implementation of Ecuador’s *Mother and Baby-Friendly Health Facilities Standard* (ESAMyN) across multiple levels of the public health system. Our findings suggest that implementing ESAMyN involved substantially more than introducing a new clinical guideline; it required organizational transformation characterized by continuous negotiation between national policy expectations and local health system realities. Across facilities, implementation was facilitated by committed leadership, continuous capacity building, local adaptation, and collaboration across institutional levels. At the same time, progress was constrained by resistance to change, workforce turnover, resource limitations, and the challenge of sustaining improvements beyond the certification process. Together, these findings highlight that implementing quality improvement initiatives in maternal and newborn health is a dynamic organizational process that depends not only on technical guidance but also on institutional capacity, adaptive leadership, and sustained system support (World Health Organization, 2016; Yohannes et al., 2026).

### Organizational change as the foundation for implementation

Our findings suggest that implementing ESAMyN represented an organizational change process rather than the simple adoption of clinical recommendations. Participants consistently described implementation as requiring changes in professional attitudes, institutional culture, and routine clinical practices, particularly those related to respectful maternity care and family participation during childbirth. These findings are consistent with implementation science frameworks, which emphasize that the successful adoption of complex healthcare interventions depends not only on the characteristics of the intervention itself but also on the organizational context, leadership, and readiness for change (Dadich et al., 2021; Pramono et al., 2025). Rather than occurring through a linear process, implementation evolved through continuous negotiation among healthcare providers, managers, and policymakers, reflecting the adaptive nature of health system change (World Health Organization, 2025c; Zarbiv et al., 2025).

Leadership emerged as a critical facilitator in navigating this transition. Hospital managers, quality improvement teams, and ESAMyN focal points not only coordinated implementation activities but also addressed resistance, fostered multidisciplinary collaboration, and reinforced institutional commitment. Similar observations have been reported in evaluations of the Baby-Friendly Hospital Initiative and other maternal and newborn quality improvement programmes, where visible leadership, shared governance, and institutional ownership have consistently been associated with greater implementation success and sustainability (Fair et al., 2024; Walsh et al., 2023). Our findings extend this evidence by showing that leadership was particularly important in supporting changes to deeply rooted professional norms and organizational routines within a middle-income health system (Bueno et al., 2023).

### Health system capacity influenced the consistency of implementation

Although ESAMyN established a standardized framework for improving maternal and newborn care, its implementation depended heavily on the capacity of health facilities to maintain trained personnel, provide supportive supervision, and allocate sufficient resources over time. Participants consistently identified continuous training as essential for reinforcing clinical competencies and maintaining adherence to the standard, particularly in settings with frequent staff turnover. However, the benefits of training were often undermined by the rotation of healthcare professionals, competing clinical demands, and limited opportunities for refresher training (Mäkelä et al., 2024; Onuorah et al., 2026). These challenges have also been reported in evaluations of the Baby-Friendly Hospital Initiative and other quality improvement programmes, where workforce instability has been identified as a major barrier to sustaining evidence-based practices (Leahy, 2025; Mäkelä et al., 2022).

Beyond human resources, participants described how shortages of infrastructure, equipment, and operational funding affected the pace and consistency of implementation across facilities (Wojcieszek et al., 2023). These findings reinforce broader evidence that the effectiveness of quality improvement initiatives depends not only on provider knowledge and motivation but also on the strength of the health system in which they are implemented (Pramono et al., 2025). The WHO Health Systems Framework highlights that sustained improvements in quality of care require coordinated investments across multiple system components, including the health workforce, financing, service delivery, governance, and essential resources (World Health Organization, 2025a). Our findings suggest that, even when national policies provide clear technical guidance, health facilities require ongoing institutional support to translate these standards into routine practice.

### Local adaptation enabled implementation but should not replace institutional support

One of the most distinctive findings of this study was the extent to which health facilities adapted the implementation of ESAMyN to overcome structural constraints. Participants described mobilizing community partnerships, reallocating local resources, organizing fundraising activities, and relying on personal commitment to compensate for shortages in infrastructure, equipment, and operational funding. These adaptive strategies allowed implementation to continue despite limited institutional resources and reflected a strong sense of ownership among healthcare teams (Pramono et al., 2025). Similar forms of local adaptation have been described in implementation research as essential mechanisms through which complex interventions are integrated into diverse organizational contexts while preserving their core objectives (Pérez-Escamilla et al., 2016; Santraine et al., 2025).

However, our findings also suggest that these adaptations came at a cost. In several facilities, implementation depended on extraordinary individual commitment, voluntary efforts, and, in some cases, personal financial contributions from healthcare workers. While these actions demonstrated remarkable professional dedication, they also exposed the vulnerability of relying on informal solutions to sustain national quality improvement initiatives. Previous studies have emphasized that adaptation can enhance implementation by increasing the contextual fit of an intervention, but excessive dependence on local improvisation may also exacerbate inequities between facilities and threaten long-term sustainability (Arslanian et al., 2022). Our findings therefore highlight the importance of balancing local flexibility with consistent institutional investment, ensuring that innovation complements, rather than substitutes for, adequate financing, infrastructure, and governance.

### Sustaining quality improvement requires institutionalization beyond certification

Our findings suggest that achieving ESAMyN certification represented an important milestone rather than the endpoint of implementation. Participants consistently emphasized that maintaining the standard required continuous monitoring, ongoing training, supportive leadership, and periodic reinforcement of recommended practices. Without these mechanisms, improvements were perceived as vulnerable to staff turnover, changing institutional priorities, and resource constraints. These findings are consistent with previous research demonstrating that the sustainability of quality improvement initiatives depends less on the initial implementation effort than on the extent to which new practices become embedded within routine organizational processes and institutional culture (Dadich et al., 2021; Zhu et al., 2025).

The experiences described by participants also underscore the importance of viewing sustainability as a system-level responsibility rather than solely the responsibility of individual health facilities. Although local leadership and professional commitment were essential drivers of implementation, long-term sustainability requires stable governance structures, dedicated financing, supportive supervision, and mechanisms for continuous quality improvement. Embedding ESAMyN within broader maternal and newborn health policies and existing health system performance frameworks may help reduce dependence on individual champions and promote more equitable implementation across facilities. These findings have implications beyond Ecuador, suggesting that national quality standards are more likely to achieve lasting improvements when implementation strategies combine strong local ownership with sustained institutional commitment at all levels of the health system.

This study has several strengths. To our knowledge, it is the first qualitative study to examine the implementation of Ecuador’s ESAMyN standard across multiple levels of the public health system. By including participants from the national Ministry of Health, health facility managers, ESAMyN focal points, quality coordinators, healthcare professionals, and women receiving care, the study captured diverse perspectives on implementation across primary, secondary, and tertiary healthcare settings in five provinces. This multi-level approach provided a comprehensive understanding of the organizational, contextual, and system-level factors influencing implementation. Furthermore, the use of semi-structured interviews and inductive thematic analysis enabled an in-depth exploration of participants’ experiences and facilitated the identification of common implementation patterns across different settings.

This study also has limitations. The findings reflect experiences from selected public health facilities in five provinces and may not represent implementation in all regions of Ecuador or in the private health sector. As with all qualitative research, the results are based on participants’ perceptions and experiences rather than direct observation of clinical practice or measurement of implementation outcomes. In addition, interviews were conducted during a specific stage of ESAMyN implementation; therefore, perceptions may evolve as the programme continues to mature. Nevertheless, the inclusion of multiple stakeholder groups and health system levels strengthened the credibility of the findings through triangulation of perspectives and allowed the identification of consistent themes across diverse implementation contexts.

## Conclusion

Implementing national maternal and newborn quality standards requires more than technical guidance. Sustainable implementation depends on leadership, organizational capacity, local adaptation, and continued institutional support. While ESAMyN demonstrates the feasibility of implementing a comprehensive quality improvement strategy in a middle-income country, maintaining these gains will require embedding the standard within routine health system governance, financing, and continuous quality improvement processes. These lessons may inform the implementation of similar maternal and newborn quality initiatives in other resource-constrained settings.

## Data Availability

The datasets analyzed during the current study are not publicly available because they are part of an institutional programme systematization and contain confidential qualitative data. Requests for access should be directed to the corresponding author and will be considered in accordance with institutional policies and applicable regulations.

## Notes

### Competing Interest Statement

The authors have declared no competing interest.

### Author Declarations

The interviews were originally conducted for programme systematization rather than research purposes. This study involved the secondary analysis of anonymized interview transcripts. According to Ecuadorian Ministerial Agreement No. 00005-2022 (Article 43), secondary analysis of anonymized data is classified as research without risk and does not require review or approval by a Research Ethics Committee.

